# Relationships between arts participation, social cohesion, and wellbeing: An integrative review and conceptual model

**DOI:** 10.1101/2024.05.01.24306077

**Authors:** Jill Sonke, Virginia Pesata, Aaron Colverson, Jane Morgan-Daniel, Alexandra K. Rodriguez, Gray Davidson Carroll, Shanaé Burch, Abel Abraham, Seher Akram, Stefany Marjani, Cassandra Belden, Hiba Karim

## Abstract

Arts and cultural strategies have increasingly been engaged by the public health sector to enhance social cohesion, health, and wellbeing, as well as to address the significant health risks posed by social isolation and loneliness. While increasing studies document relationships between arts participation, social cohesion, and wellbeing uniquely, few studies have investigated the relationships between all three and, to date, no evidence synthesis has been conducted on this topic. To address this gap, this integrative review aimed to identify, describe, and synthesize research on arts participation, social cohesion, and wellbeing in a community context by addressing the question: what is the evidence base regarding relationships between arts participation, social cohesion, and well-being? Literature searches were conducted using 10 databases, and 18 articles met inclusion criteria – 16 original research articles and two reviews. Results provide insights on modes and forms of arts participation used, and offer four themes that articulate dimensions of and relationships between arts participation, social cohesion, and well-being, as distilled from the studies. Further, this review offers a conceptual model derived from these themes. The model depicts the relationships found between these concepts in the articles and highlights specific components of these relationships that may help to guide future practice, research, and policy that seeks to leverage the power of the arts to build social cohesion and wellbeing in communities. Prospective studies are needed to test these relationships as well as the potential role of social cohesion as a mechanism for building well-being in communities.

## Introduction

In recent years, arts and cultural strategies have been increasingly engaged by the public health sector to enhance community health and well-being, as well as to address social drivers and causes of health inequities and disparities in the United States (US) (1–4). This uptake is in part due to the growing body of evidence that links arts and cultural participation to notable impacts on health and well-being at the population level in this country, including on mental health (5), flourishing among young people (6), healthy aging (7), and even mortality (8). Additionally, as the significant health risks posed by social isolation and loneliness are better understood, arts and cultural strategies are increasingly recognized as available resources for countering isolation and building social cohesion in communities (9, 10).

The 2023 report by US Surgeon General, Dr. Vivek Murthy, highlights that half of American adults experience loneliness, and that loneliness is a serious health risk at both the individual and collective levels (11). The report notes that the health risks of loneliness are greater than that of obesity or inactivity, and calls with urgency for “a movement to mend the social fabric of our nation” (p. 5). This urgency is underscored by the isolation caused by the COVID-19 pandemic and heightened socio-political divisions in the US, which highlight the need not only for opportunities for social connection, but also for building social cohesion, defined in Healthy People 2030 as “the strength of relationships and a sense of solidarity among members of a community” (12).

The notion of social fabric referred to in the Surgeon General’s report is not just about social relationships and connections. A strong social fabric refers as well to social cohesion, or the sense of belonging to a group and a willingness to participate and share in it (13). While definitions vary greatly, many refer to the attitudes and behaviors of members of a community or group, and not simply proximity or connection (14). As such, social cohesion is critical to a community’s ability to respond to challenges and to create conditions in which its members can thrive.

Social cohesion is often thought of generally as the “glue that holds societies together” (15). The concept can be found in writings from the 14^th^ century attributed to Arab sociologist and philosopher, Ibn-Haldun who defined the term “asabiyyah” to mean group feeling or social cohesion, described as a mix of unity and group consciousness (16). Early Western theorists, including Émile Durkheim and Charles Horton Cooley, laid the groundwork for understanding social cohesion as integral to societal functioning and well-being, highlighting collective behavior and interdependence (17). More current definitions, although varied, often focus on the strength of relationships, solidarity, belonging, orientation toward a common good, and willingness to participate (12, 17–19). For example, the Council of Europe defines social cohesion “as the capacity of a society to ensure the well-being of all its members—minimizing disparities and avoiding marginalization—to manage differences and divisions and ensure the means of achieving welfare for all members” (20). At times, the term has, problematically, been used to refer to the sense of identification and emotional ties among people who share the same characteristics (21), but this notion of homogeneity as a variable in social cohesion has since been recognized to be linked to economic deprivation (22). More current definitions emphasize multiculturism and the absence of conflict across differences such as wealth, ethnicity, and race (23, 24).

Arts participation has been explored as a means for building social cohesion in US communities (25) and other regions (26, 27), as well as for enhancing the well-being of individuals and communities. The *We-Making* Theory of Change, for example, offers a framework that articulates a relationship between place-based arts and cultural strategies, social cohesion, and community well-being in US communities (28). This theory of change identifies ingredients in place-based arts and cultural strategies that amplify specific drivers of social cohesion, which in turn lead to increased equitable community well-being, including physical and mental health and civic capacity for change. This theory of change offers a promising model for guiding programming and policy that can increase utilization of available arts and cultural resources to address the critical issue of social isolation in the US communities.

While increasing studies document relationships between arts participation and both social cohesion and well-being uniquely, few studies have investigated the relationships between all three and, to date, no evidence synthesis has been conducted on this topic. To address this gap, this integrative review aimed to identify, describe, and synthesize literature that investigates arts participation, social cohesion, and well-being in a community context. It builds on work undertaken through the *We-Making* initiative (28), and also serves as a foundational study within the broader research agenda developed to assess the One Nation/One Project initiative, a national arts and health initiative designed to activate the power of the arts for social cohesion, health, and well-being and to repair the social fabric of US communities following the COVID-19 pandemic. The review was designed to address the question: *what is the evidence regarding relationships between arts participation, social cohesion, and well-being?*

## Methods

### Methodology

This study utilized an integrative review methodology (29) to examine published literature that investigates arts participation, social cohesion, and well-being. Given the specific and nascent nature of this topic, the integrative review methodology was chosen over others (e.g., scoping or systematic review) to include a broad range of evidence, including theoretical, methodological, and empirical literature. Although no formal guidelines exist for conducting integrative reviews, most align with scoping review guidelines and follow a systematic yet holistic approach to minimize bias during literature searching, screening, data extraction, and critical appraisal of included studies (29). This study’s methodology development was guided by PRISMA for Scoping Reviews and adopted a holistic approach to synthesizing empirical and theoretical literature to develop a new model or framework (29, 30).

### Guiding Definitions

The study utilized four definitions to guide its inclusion and exclusion criteria, as well as data extraction and analysis. Each of the concepts investigated in this study evades a single, widely accepted definition. Therefore, the study utilized definitions that offered both breadth and specificity in alignment with the nature of the study’s focus.

Community was defined as a group inhabiting a common territory or having one or more common ties (31). Within this definition, we recognized communities of people sharing common geographic areas, including those composed of culturally distinct members (people with similar cultural identities) and culturally heterogeneous groups (people of different cultural identities related to, among other things, ethnicity, language, race, values, age, or sense of place). We also recognized transient communities, including temporary, resettled, dispersed or displaced residents, including migrant, diasporic, or student communities.

The review utilized a broad and inclusive concept of “arts participation”. The search strategy included a broad range of terms representing the arts (See S1 Search Strategy: Sample Search Strategy for PubMed), and both our search strategy and inclusion criteria reflected the definition of “arts participation” developed for the purpose of public health research (32). This definition includes modes, or ways, in which people engage with the arts, and includes examples of various art forms intended to frame arts participation broadly and inclusively.

The study engaged the Healthy People 2030 definition of social cohesion (33), which refers to “the strength of relationships and a sense of solidarity among members of a community”, along with several variables for social cohesion that were developed in the project’s broader research agenda. In all, our definition of social cohesion was represented by nine variables: social relationships, social networks, solidarity, belonging, social capital, participation, trust, inclusion, and social support.

Well-being was defined in alignment with the Robert Wood Johnson Foundation definition: “the comprehensive view of how individuals and communities experience and evaluate their lives, including their physical and mental health and having the skills and opportunities to construct meaningful futures” (34). While there is no consistent cross-sector definition of well-being, subjective well-being is increasingly recognized as an important health indicator.

## Preliminary Searching and Protocol

An initial search for similar pre-existing reviews or protocols was carried out on February 25, 2022. The keyword search strategy (art OR arts OR artistic OR artist OR artists) AND (well-being OR “well-being” OR “well-being”) AND (social) was used in BioMed Central Systematic Reviews, Campbell Collaboration Education Group, Cochrane Database of Systematic Reviews, JBI Systematic Review Register, JBI Evidence Synthesis, and PROSPERO: International Prospective Register of Systematic Reviews. No reviews or protocols were located that focused on the relationship between arts participation, social cohesion, and well-being. The protocol for this study was not registered, in accordance with previous work’s guidelines (29).

### Searching and Eligibility Criteria

A health sciences librarian developed the search strategy with research team input. Test searching occurred in the database PubMed between February and September 2022. The search strategy was based on the Population, Concept, Context (PCC) framework that was used to determine the eligibility criteria for the review (35). Using the definitions noted above, the population was community, the concepts were arts participation and social cohesion, and the context was well-being.

The final literature searches were conducted between November 3-8, 2022, using subject headings and keywords in 10 databases, which were all chosen for their topic focus on arts and health. The databases searched were EBSCOhost’s Alt HealthWatch, Art and Architecture Source, CINAHL, PsycINFO, Psychology and Behavioral Sciences Collection; ProQuest’s Applied Social Sciences Index and Abstracts, Performing Arts Periodicals Database; PubMed; Scopus (Elsevier); and Web of Science (Clarivate Analytics). The database searches included published and grey literature. The only search limits used were English language and 2000 onwards for the date. A sample search strategy is provided as supporting information (See S1 Search Strategy). The reference lists of all included studies were checked for additional articles on September 7, 2023.

To increase the breadth of the literature, this review included all research designs, program evaluation reports, systematic and scoping reviews, and doctoral dissertations. Grey literature was searched and included articles that fit the inclusion criteria and were deemed credible through agreement of the research team. These included articles from scientific journals, as well as reports from professional organizations, governmental, and non-governmental organizations, or agencies (i.e., the World Health Organization). Additionally, the project team hand-searched the following web archives and databases: National Organization for Arts in Health (NOAH), Alliance for the Arts in Research Universities (a2ru), American Art Therapy Association, American Music Therapy Association, the University of Florida Center for Arts in Medicine Research Database, The Wallace Foundation, The International Expressive Art Association, University College London, and the National Endowment for the Arts.

### Selection of Evidence

Title, abstract, and full-text screening were undertaken by independent blinded pairs of researchers utilizing the inclusion criteria presented in Table 1 in Covidence. Agreement of two reviewers was required with a third reviewer resolving differences, as needed.

**Table 1.** Inclusion criteria.

| Inclusion Criteria |  |
| --- | --- |
| Overall | 1. Human Studies |
| Intervention | Studies of arts participation as an intervention addressing social cohesion (as a mechanism or outcome) and well-being (as an outcome). |
| Outcomes | Outcomes related to social cohesion and well-being, as defined for this review, OR outcomes related to well-being with social cohesion as a mechanism. |
| Timeframe | Published during or after the year 2000. |
| Language | Written in English language. |
| Sources | The source was from a peer-review journal or other credible source including universities, professional organizations, governmental, and global organizations (i.e. the World Health Organization) and nongovernmental organizations (NGO). |
| Measurement/Research Methods | 1. Evidence of defined measures used to arrive at outcomes.<br>2. Qualitative, quantitative, mixed methods, etc. |

The review emphasized inclusion of all three concepts - arts participation, social cohesion, and well-being - allowing for social cohesion to be present as either a mechanism or an outcome. Articles that presented relationships between only two of the three concepts were excluded.

### Quality Assessment

After selection, quality assessment was conducted by two researchers using the Mixed Methods Appraisal Tool (MMAT) Version 2018 (36), with discrepancies resolved by a third reviewer until consensus was achieved. Following quality assessment, the research team discussed the selected articles together to ensure agreement around the concepts of arts participation, social cohesion, and well-being to ensure that the sources met the inclusion criteria.

### Data Extraction

Using a screening form developed in Covidence’s data extraction tool, the research team extracted the data. It was then checked for accuracy and completeness by a separate team member. Data were extracted to examine numerous dimensions of the studies, as presented in the results section below.

### Data Analysis

A mixed-methods approach was used for describing and developing inferences related to the relationships between arts participation, social cohesion, and well-being from across the included studies. The analysis included quantitative analyses of study designs, populations studied, unit of social cohesion analysis (within-group or broader community cohesion), modes of arts participation, and forms of art utilized in intervention programs. A qualitative and quantitative context-mechanism-outcomes approach was used to examine whether social cohesion was present as a mechanism or outcome or both.

The papers were also coded for dimensions, or components, of social cohesion that were reported as results (as either mechanisms, outcomes, or both). A directed approach was used to code for components of the Healthy People 2030 definition of social cohesion, along with the variables of social cohesion used in the One Nation/One Project initiative’s overarching research agenda. The definition and variables together included nine components: social support, inclusion, trust, participation, social capital, belonging, solidarity, social relationships, and social networks.

Finally, a directed qualitative thematic content analysis was used to develop inferences related to the review’s research question: *what is the evidence base regarding relationships between arts participation, social cohesion, and well-being?*

## Results

### Numerical Summary

As shown in Table 1, the database search yielded 3156 results; hand-researching the reference lists of the included studies led to 88 additional references added to Covidence, in addition to 2 references from grey literature. Covidence identified 1433 duplicates, whereas 7 additional duplicates were identified manually. The final number of title/abstracts screened was therefore 1806, with 1454 marked as irrelevant. Full-text studies (n=352) were assessed for eligibility, with 334 excluded.

After the title, abstract and full text review, 18 studies were included in the final review (See Table 2). Of these, there were 2 reviews, 1 doctoral dissertation, and 2 reports. The remaining 13 records were original research articles.

**Figure 1.** PRISMA Diagram.

**Table 2.**
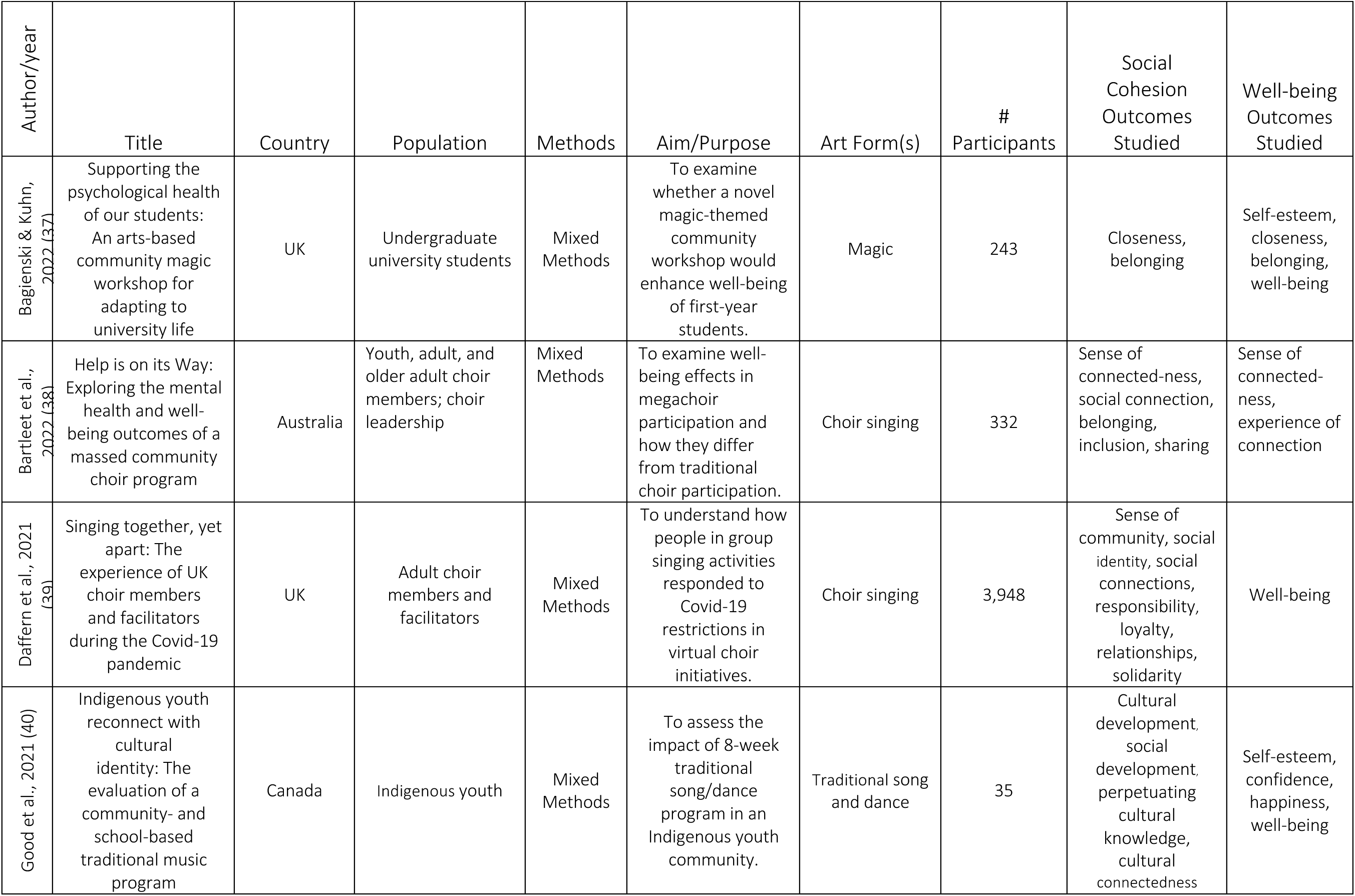
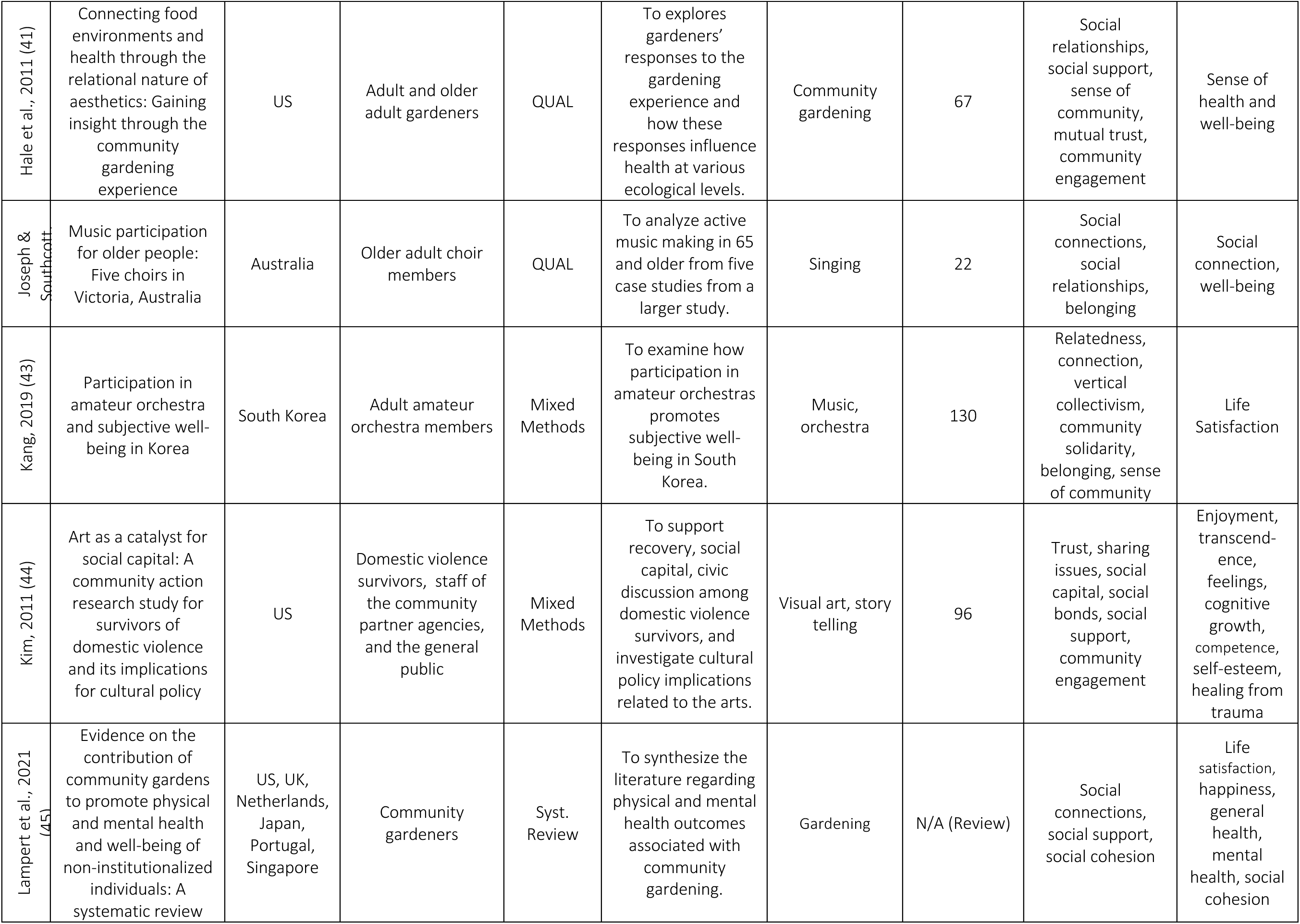
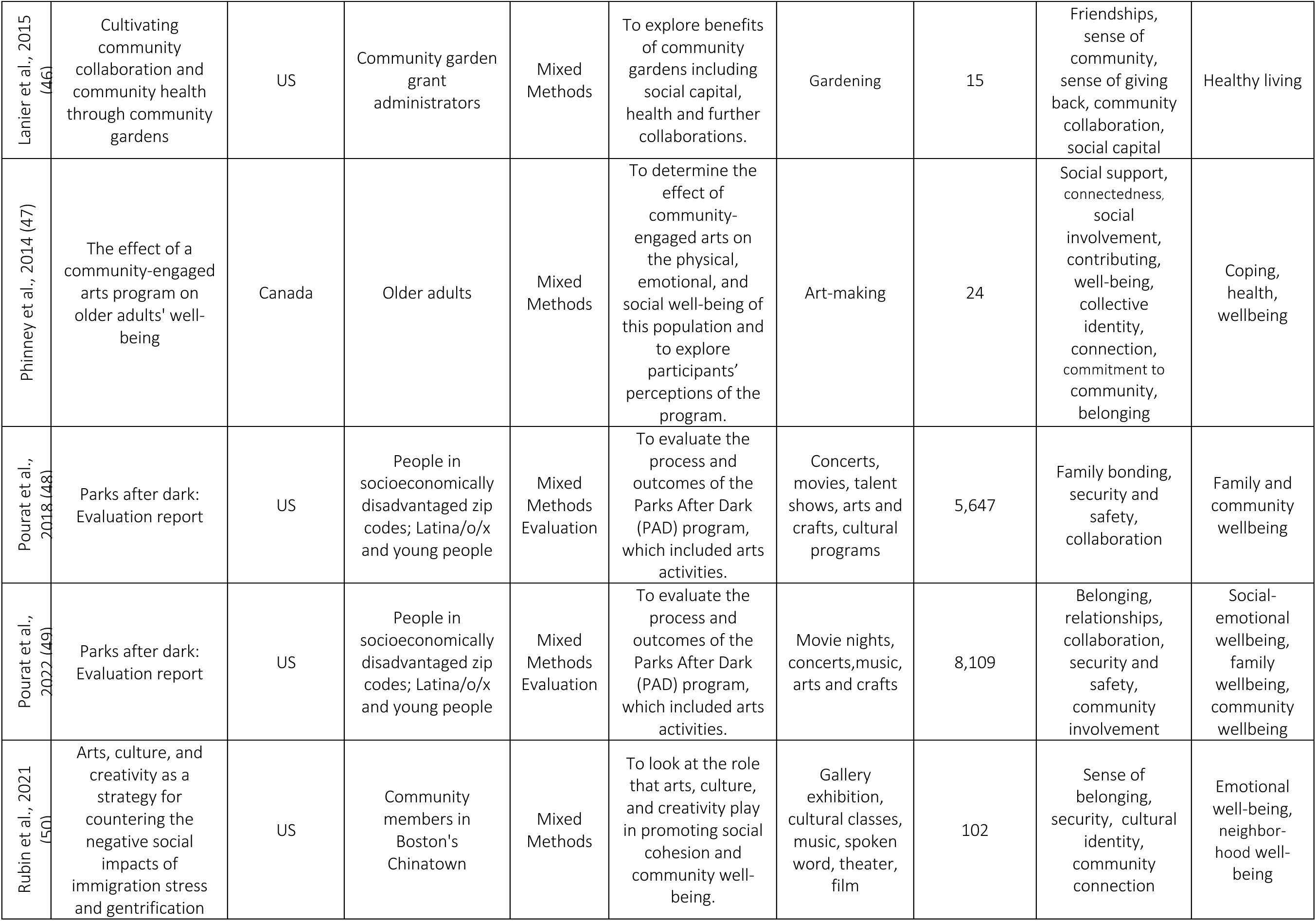
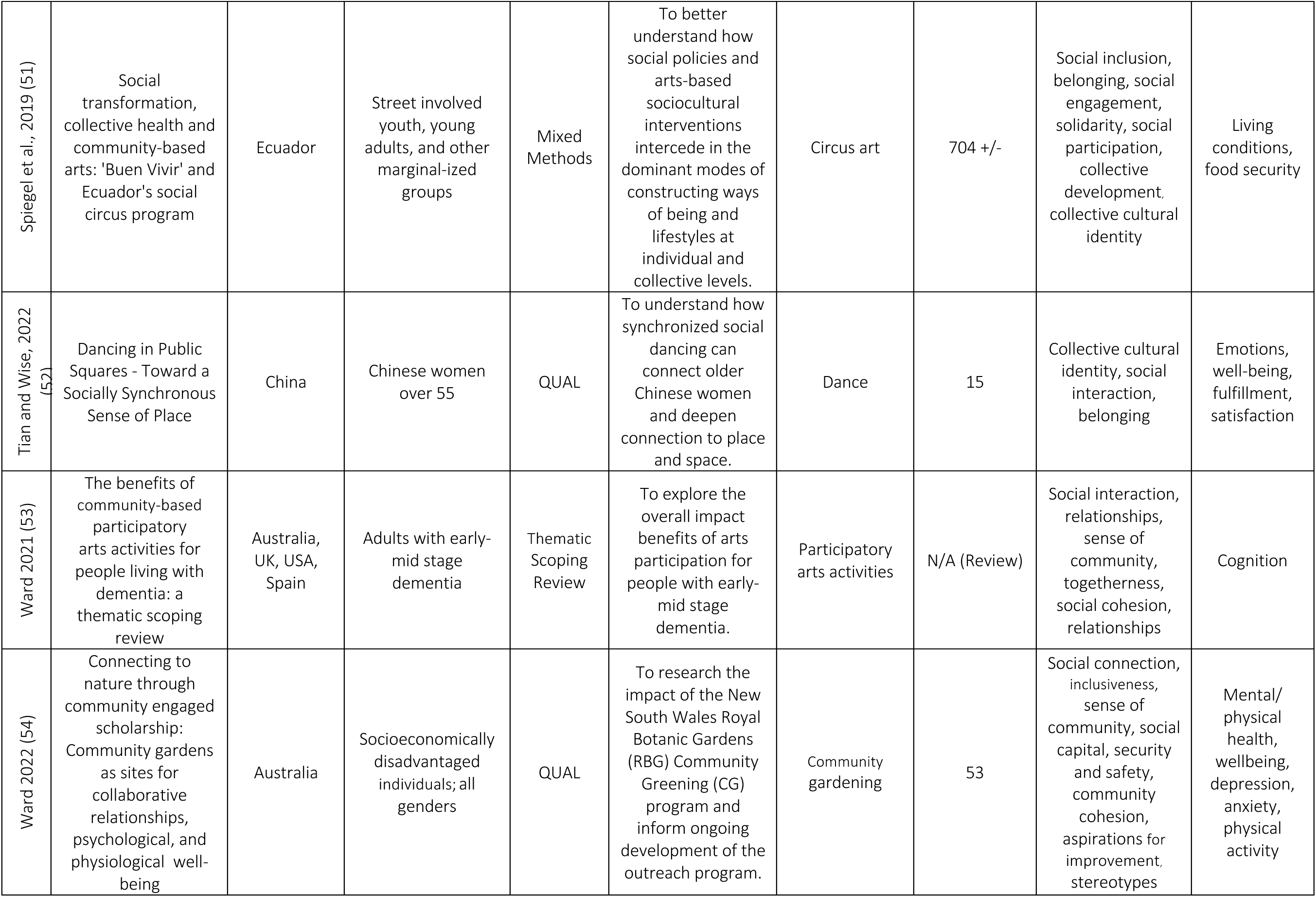
Included Articles: Key Data Extracted.

| Author/year | Title | Country | Population | Methods | Aim/Purpose | Art Form(s) | # Participants | Social Cohesion Outcomes Studied | Well-being Outcomes Studied |
| --- | --- | --- | --- | --- | --- | --- | --- | --- | --- |
| Bagiński & Kuhn, 2022 (37) | Supporting the psychological health of our students: An arts-based community magic workshop for adapting to university life | UK | Undergraduate university students | Mixed Methods | To examine whether a novel magic-themed community workshop would enhance well-being of first-year students. | Magic | 243 | Closeness, belonging | Self-esteem, closeness, belonging, well-being |
| Bartleet et al., 2022 (38) | Help is on its Way: Exploring the mental health and well-being outcomes of a massed community choir program | Australia | Youth, adult, and older adult choir members; choir leadership | Mixed Methods | To examine well-being effects in megachoir participation and how they differ from traditional choir participation. | Choir singing | 332 | Sense of connected-ness, social connection, belonging, inclusion, sharing | Sense of connected-ness, experience of connection |
| Daffern et al., 2021 (39) | Singing together, yet apart: The experience of UK choir members and facilitators during the Covid-19 pandemic | UK | Adult choir members and facilitators | Mixed Methods | To understand how people in group singing activities responded to Covid-19 restrictions in virtual choir initiatives. | Choir singing | 3,948 | Sense of community, social identity, social connections, responsibility, loyalty, relationships, solidarity | Well-being |
| Good et al., 2021 (40) | Indigenous youth reconnect with cultural identity: The evaluation of a community- and school-based traditional music program | Canada | Indigenous youth | Mixed Methods | To assess the impact of 8-week traditional song/dance program in an Indigenous youth community. | Traditional song and dance | 35 | Cultural development, social development, perpetuating cultural knowledge, cultural connectedness | Self-esteem, confidence, happiness, well-being |
| Hale et al., 2011 (41) | Connecting food environments and health through the relational nature of aesthetics: Gaining insight through the community gardening experience | US | Adult and older adult gardeners | QUAL | To explore gardeners' responses to the gardening experience and how these responses influence health at various ecological levels. | Community gardening | 67 | Social relationships, social support, sense of community, mutual trust, community engagement | Sense of health and well-being |
| Joseph & Southcott | Music participation for older people: Five choirs in Victoria, Australia | Australia | Older adult choir members | QUAL | To analyze active music making in 65 and older from five case studies from a larger study. | Singing | 22 | Social connections, social relationships, belonging | Social connection, well-being |
| Kang, 2019 (43) | Participation in amateur orchestra and subjective well-being in Korea | South Korea | Adult amateur orchestra members | Mixed Methods | To examine how participation in amateur orchestras promotes subjective well-being in South Korea. | Music, orchestra | 130 | Relatedness, connection, vertical collectivism, community solidarity, belonging, sense of community | Life Satisfaction |
| Kim, 2011 (44) | Art as a catalyst for social capital: A community action research study for survivors of domestic violence and its implications for cultural policy | US | Domestic violence survivors, staff of the community partner agencies, and the general public | Mixed Methods | To support recovery, social capital, civic discussion among domestic violence survivors, and investigate cultural policy implications related to the arts. | Visual art, story telling | 96 | Trust, sharing issues, social capital, social bonds, social support, community engagement | Enjoyment, transcendence, feelings, cognitive growth, competence, self-esteem, healing from trauma |
| Lampert et al., 2021 (45) | Evidence on the contribution of community gardens to promote physical and mental health and well-being of non-institutionalized individuals: A systematic review | US, UK, Netherlands, Japan, Portugal, Singapore | Community gardeners | Syst. Review | To synthesize the literature regarding physical and mental health outcomes associated with community gardening. | Gardening | N/A (Review) | Social connections, social support, social cohesion | Life satisfaction, happiness, general health, mental health, social cohesion |
| Lanier et al., 2015<br>(46) | Cultivating community collaboration and community health through community gardens | US | Community garden grant administrators | Mixed Methods | To explore benefits of community gardens including social capital, health and further collaborations. | Gardening | 15 | Friendships, sense of community, sense of giving back, community collaboration, social capital | Healthy living |
| Phinney et al., 2014 (47) | The effect of a community-engaged arts program on older adults' well-being | Canada | Older adults | Mixed Methods | To determine the effect of community-engaged arts on the physical, emotional, and social well-being of this population and to explore participants' perceptions of the program. | Art-making | 24 | Social support, connectedness, social involvement, contributing, well-being, collective identity, connection, commitment to community, belonging | Coping, health, wellbeing |
| Pourat et al., 2018 (48) | Parks after dark: Evaluation report | US | People in socioeconomically disadvantaged zip codes; Latina/o/x and young people | Mixed Methods Evaluation | To evaluate the process and outcomes of the Parks After Dark (PAD) program, which included arts activities. | Concerts, movies, talent shows, arts and crafts, cultural programs | 5,647 | Family bonding, security and safety, collaboration | Family and community wellbeing |
| Pourat et al., 2022 (49) | Parks after dark: Evaluation report | US | People in socioeconomically disadvantaged zip codes; Latina/o/x and young people | Mixed Methods Evaluation | To evaluate the process and outcomes of the Parks After Dark (PAD) program, which included arts activities. | Movie nights, concerts, music, arts and crafts | 8,109 | Belonging, relationships, collaboration, security and safety, community involvement | Social-emotional wellbeing, family wellbeing, community wellbeing |
| Rubin et al., 2021<br>(50) | Arts, culture, and creativity as a strategy for countering the negative social impacts of immigration stress and gentrification | US | Community members in Boston's Chinatown | Mixed Methods | To look at the role that arts, culture, and creativity play in promoting social cohesion and community well-being. | Gallery exhibition, cultural classes, music, spoken word, theater, film | 102 | Sense of belonging, security, cultural identity, community connection | Emotional well-being, neighborhood well-being |
| Spiegel et al., 2019 (51) | Social transformation, collective health and community-based arts: 'Buen Vivir' and Ecuador's social circus program | Ecuador | Street involved youth, young adults, and other marginal-ized groups | Mixed Methods | To better understand how social policies and arts-based sociocultural interventions intercede in the dominant modes of constructing ways of being and lifestyles at individual and collective levels. | Circus art | 704 +/- | Social inclusion, belonging, social engagement, solidarity, social participation, collective development, collective cultural identity | Living conditions, food security |
| Tian and Wise, 2022 (52) | Dancing in Public Squares - Toward a Socially Synchronous Sense of Place | China | Chinese women over 55 | QUAL | To understand how synchronized social dancing can connect older Chinese women and deepen connection to place and space. | Dance | 15 | Collective cultural identity, social interaction, belonging | Emotions, well-being, fulfillment, satisfaction |
| Ward 2021 (53) | The benefits of community-based participatory arts activities for people living with dementia: a thematic scoping review | Australia, UK, USA, Spain | Adults with early-mid stage dementia | Thematic Scoping Review | To explore the overall impact benefits of arts participation for people with early-mid stage dementia. | Participatory arts activities | N/A (Review) | Social interaction, relationships, sense of community, togetherness, social cohesion, relationships | Cognition |
| Ward 2022 (54) | Connecting to nature through community engaged scholarship: Community gardens as sites for collaborative relationships, psychological, and physiological well-being | Australia | Socioeconomically disadvantaged individuals; all genders | QUAL | To research the impact of the New South Wales Royal Botanic Gardens (RBG) Community Greening (CG) program and inform ongoing development of the outreach program. | Community gardening | 53 | Social connection, inclusiveness, sense of community, social capital, security and safety, community cohesion, aspirations for improvement, stereotypes | Mental/ physical health, wellbeing, depression, anxiety, physical activity |

There is no PRISMA checklist developed for integrative reviews, for this reason the PRISMA ScR checklist was used. (See Supplemental materials S 2). The Mixed Methods Appraisal Tool (38) was used to conduct quality appraisal for original research articles (N=13) and one dissertation (total N=14). Two evaluation reports (48,49) and two reviews (45,53) were excluded from quality appraisal. The (MMAT) criteria is as follows: 1) was/is there clear research question(s); and 2) did the collected data address the research question(s)? Additionally, the sources were evaluated based on their study design (e.g., qualitative, quantitative – randomized or non-randomized, or descriptive - and mixed methods) with the MMAT methodological quality criteria. All the research sources passed the quality appraisal.

### Study Designs

Most of the included articles (n=11) used mixed methods in their research or evaluation, while some used qualitative (n=4) or quantitative (n=1) methods alone, and two were review articles. One of the review articles was a systematic review, and the other was a thematic scoping review. The research articles (n=14) and evaluation reports (N=2) used a range of data collection methods including surveys or questionnaires in eleven articles, and interviews and/or focus groups in mixed methods and qualitative studies.

### Populations Studied

Twelve countries were represented in the review. The 16 research and evaluation articles presented findings from programs in the United States (n=6), China (n=1), Canada (n=2), the United Kingdom (n=2), Australia (n=3), South Korea (n=1) and Ecuador (n=1). Both review articles presented findings from numerous countries (See Table 2). Populations studied in the included articles varied across age, gender, and other demographics. While most articles included all gender identities, three focused solely on people who identified as women. Adults were the most commonly studied age group across the articles.

Collectively, the 16 research and evaluation articles included 11,621 total participants. Most participants (n=6,669) were part of choir groups. Among the articles that reported on the gender of the participants (n= 3,708), 75% (n=2784) identified as female and 25% (n=924) identified as male. Other population groups represented in the studies included public housing residents, women, migrant communities, college students, domestic violence survivors, Indigenous youth, people living with dementia, youth living in poverty, and older adults.

### Modes of Arts Participation

Modes of arts participation (see Table 3) were assessed in alignment with the review’s definition of arts participation (32). All of the 16 research and evaluation articles studied programs that included creating, practicing, performing, and sharing art; and multiple modes were reported in some studies.

**Table 3.** Modes of Participation.

| Mode | n | % |
| --- | --- | --- |
| Attending live arts and cultural events and activities | 2 | 10 |
| Creating, practicing, performing, and sharing art | 18 | 100% |
| Participating in social, civic, spiritual, and cultural arts practices | 5 | 26% |
| Consuming arts via electronic, digital, or print media | 0 | 0 |
| Learning in, through, and about the arts | 3 | 16% |

Choir programs were particularly common in this category. For example, Bartleet (2022) assessed the experiences of 305 choir members in Australia and reported that sharing, belonging, and inclusion promoted positive well-being outcomes (38). The perspectives of socially distanced choirs (due to COVID-19) were investigated by Daffern (2021) using a cross-sectional online survey with the input of 3,948 choir members in the UK (39). A thematic scoping review focusing on mixed-art forms reported that community-based arts activities have positive impacts and holistic benefits to individuals with early and moderate stages of dementia (53). This study noted that active participation and interaction were integral parts of the success of the interventions.

Also common among modes of participation were social, civic, spiritual, and cultural practices. For example, Good (2021) reported that a community- and school-based participatory music program gave students the opportunity to connect with each other and their community and to work together toward a common goal, and that the program enhanced well-being among participants (40).

### Art Forms

Among the included articles, there was an equal representation of programs that included the categories of: 1) music; 2) visual art, crafts, and design; and 3) multiple arts forms (see Figure 2).

**Figure 2.** Art Forms.

The four programs included in the visual arts, crafts and design category were all gardening programs. The multiple art forms category included visual arts and craft, music, dance, theatre, literary arts, spoken word, film, concerts and talent shows, and cultural programs. For example, Phinney (2014), studied the participation of older adults in weekly workshops offered over a three-year period at community centers (47). The workshops included engagement in various visual and performing art forms that led to co-creation of performances and art pieces. The Parks After Dark program in an urban US community included concerts, movies, talent shows, arts and crafts, cultural programs (48). In its evaluation, alongside recreation, sports, clubs and other activities, these arts and entertainment programs were rated as the most popular among participating community members.

### Unit of Analysis: Communities and/or Activity Groups

In 11 of the 16 research and evaluation articles and in the two reviews, social cohesion was assessed in relation to a geographic community in which participants lived, including neighborhoods, cities, and entire countries. For example, Hale and colleagues (2011) collected data from community gardeners not just in one garden, but from across an entire urban US city, and Bartleet (2022) did so from choir members across Australia (38, 41). However, in five studies social cohesion was considered in relation to the intervention or program group as a community. These articles were included because of the relevance or applicability of the findings to social cohesion in geographic communities. Additionally, in some cases, such as in Kang (2019), the program communities were large and created communities that aligned with the review’s definition of community (43).

### Components of Social Cohesion

The nine components of social cohesion included in the review’s guiding definition - social support, inclusion, trust, participation, social capital, belonging, solidarity, social relationships, and social networks - were each measured in one or more articles. Related concepts were coded into these nine component categories, the most common variable being social relationships, often reported as social connections. Figure 3 presents the frequency of codes for each of the nine components.

**Figure 3.** Components of Social Cohesion.

### Social Cohesion as a Mechanism

This study was interested in whether social cohesion was found to be a mechanism to well-being, or an outcome alongside well-being. Among the 16 original research articles, social cohesion was reported as a mechanism or potential mechanism linked to well-being in eight articles (38, 39, 41, 42, 47, 51, 52, 54). For example, in a study of a massed choir population across Australia, the authors suggested that social cohesion, specifically social connections and sense of belonging, can be fostered through choir participation and in turn, can contribute to enhanced mental health and well-being (38). Theme 4 below provides more examples.

### Themes

The directed qualitative thematic content analysis of included articles was directed by the review’s research question and undertaken through the lens of its guiding definitions as well as the We-Making Framework (28). It sought to identify how, collectively, the included articles articulate relationships between arts participation, social cohesion, and well-being. Four themes were developed.

#### Theme 1. Arts participation in community spaces that includes creative physical or tactile engagement can build social cohesion

Four articles highlighted the significance of engagement in community spaces for facilitating social relationships, solidarity, belonging, and social capital. One study reported that utilizing public squares in China for dancing fostered a shared sense of belonging to populations and cultures (52). Physical creative activity within these public squares cultivated relationships between identity and the historical context of the spaces, which was positively correlated with emotions and social cohesion. Participants in this study reported that through their interactions they had increased satisfaction and fulfillment and, in turn, increased well-being.

Kim (2011) reported on the use of community spaces for art-making workshops for female-identifying people who had experienced domestic violence (44). The program created a public visual gallery for storytelling and to facilitate dialogue and foster social capital. As a result of this collaboration, participants developed a sense of trust with others that cultivated social cohesion and personal well-being. While most studies found that arts participation in community spaces fostered social cohesion, Daffern (2021) found that choirs conducted virtually during the COVID-19 pandemic negatively affected the well-being of some participants, reporting that, “the virtual choir in itself actually had a strong negative impact on well-being” (p. 8) (39). The participants reported that singing alone and virtually increased stress, anxiety, and loneliness. However, many participants also found value and connection in the remote access.

#### Theme 2: Culturally rooted arts programs can enhance individual and shared cultural identity, belonging, solidarity, and cooperation

This theme highlights that community-based arts participation that is rooted in cultural identities, traditions, and practices may be a particularly effective means for engaging people in shared activities that can build important dimensions of social cohesion. For example, in a study of how a community arts center can serve as ethnic enclaves that build social cohesion and well-being, Rubin and colleagues (2021) found that the center and its creative activities played a significant role in maintaining social cohesion despite the threat of gentrification and cultural displacement (50). The study underscored the importance of the art center in fostering a sense of security, belonging, and shared cultural identity, and in maintaining social cohesion. Culturally rooted arts program can also facilitate the process of reclaiming cultural traditions as a component of intergenerational healing. Good et al. (2021) reported on how Indigenous youth connected with their heritage through song and dance, sewing regalia, making drums, and learning native language lyrics in a school-based traditional music program (40). The program documented concurrent improvements in personal, cultural, and social development, while generating connection to cultural identity and a strengthened sense of community.

This theme also brings to light ways that arts participation creates spaces for cross-cultural exchanges that can build social cohesion among people of diverse cultural backgrounds. For example, Joseph & Southscott’s (2018) research found that, “choirs formed by culturally and linguistically diverse groups provide opportunities through performance for the maintenance and transmission of heritage to family, friends and wider community” (p. 187) (42).

#### Theme 3: Co-creation and social relationships cultivate commitment to a group and program, which can result in more regular participation, and in turn, enhance benefits

Several articles highlighted that co-creation, such as community members working together to create a performance or exhibit, can cultivate a commitment to fellow program participants and to a program, resulting in more regular or sustained participation. In turn, more regular participation was shown to result in more benefits to social cohesion, health, and well-being. For example, in a study of a national circus program including five community participatory circus programs in Ecuador Spiegel et al. (2019) found that participants in a community circus project experienced an enhanced sense of group community bonding and belonging and, as a result, their engagement in the community and well-being grew (51). They also highlighted ways in which social inclusion and collective practice are intrinsically linked and mutually affect one another.

In a study of in a community orchestra group, Kang (2019) found that “In a collaborative process of playing in an orchestra, members shared responsibility and resources, and communicated and negotiated among members to achieve the communal goal” (p. 10) (43).

They also found enhanced subjective well-being among participants to be associated with a sense of solidarity and community belonging that is cultivated through participation in the orchestra, noting that orchestras can create communities in which different cultural values can be harmonized, which can be an important contributor to social cohesion. Similarly, Phinney and Moody (2014) found that participants in co-creation of visual arts exhibits and performances felt a sense of connection and commitment to each other, a stronger sense of belonging to the larger community, and that in turn this commitment to one another and the program drew participants back consistently (47). Ward (2022) reported that community gardening changed participants’ satisfaction with being a part of a community and enhanced both community participation and people’s ability to make contributions to the community (54). These feelings contributed to a commitment to gardening, which in turn enabled enhancements in health, well-being, and social capital.

#### Theme 4: Social cohesion, and its various components, may serve as a mechanism to enhancing well-being

As noted above, six articles presented studies that suggest that social cohesion can act as a mechanism to well-being in communities. Spiegel et al. (2019) found that creative programs engaged to promote social transformation (through social inclusion, social engagement, culture-sharing, and collective practice) can contribute to both individual and collective well-being (51). Joseph and Southcott (2018) reported that choir members experienced a profound feeling of social connection, which overcame social isolation and positively impacted their sense of well-being (p. 187) (42). Phinney and Moody (2014) also found that the social cohesion that was cultivated through arts participation and Co-creation generated improvements in physical and social health and well-being (47).

Similarly, Ward et al. (2022) reported findings of a study of a community gardening program in New South Wales that community gardening supported social cohesion (social connection, inclusiveness, and a sense of community), which in turn enhanced well-being along with health and social capital (54). Finally, Tian and Wise (2022) studied a public square dance program for women over age 55, most of whom reported experiencing loneliness, in a suburban Chinese community (52). They found that synchronous group dancing in the public square built numerous dimensions of social cohesion, including being a part of a group, which led to enhanced sense of place, socialization, and well-being.

### Conceptual Model

Figure 4 below presents a conceptual model representing the relationships between arts participation, social cohesion, and well-being articulated in the included articles. The model depicts the relationships found between these concepts in the articles, and also highlights specific components of these relationships.

**Figure 4.** Conceptual Model of Relationships between Arts Participation, Social Cohesion, and Well-being in the Reviewed Articles.

The model suggests a cycle in which community-based arts participation, particularly wherein co-creation - which refers to activities such as co-crafting and preparing for a performance or art exhibit - and/or cultural sharing are involved, may facilitate relationships and group belonging. Additionally, through both co-creation and cultural sharing (arts activities centered in cultural identities, practices, and traditions) participants build relationships, a sense of belonging to a community, and solidarity. This in turn can lead to a greater commitment to the program and fellow participants, which may lead to more regular participation in the program. This cycle of arts participation can build social cohesion and (potentially, in turn) enhance both individual and community well-being. It is important to note that while in some cases social cohesion was suggested to act as an important mechanism for enhancing well-being, arts participation itself also yielded direct benefits to both well-being and health.

## Discussion

This study was undertaken as a foundational study in the research agenda for the national One Nation/One Project initiative. Recognizing a need for place-based strategies for rebuilding the social fabrics and well-being of communities disrupted by the COVID-19 pandemic, this integrative review aimed to identify, synthesize, and describe literature that investigates arts participation, social cohesion, and well-being in a community context in the US. It did not seek to synthesize the abundance of literature that has previously associated arts participation with well-being (5, 55), or solely with social cohesion (25). It sought specifically to consider studies that included all three constructs.

The review synthesized the findings of 18 articles - 16 original research articles and two evidence reviews. It describes this literature by quantifying significant characteristics across the studies and presents four themes that describe the relationships that emerged from these articles between arts participation, social cohesion, and well-being. These findings highlight key dimensions of community-based arts participation that may contribute to building both social cohesion and well-being. They also support the need for further study of social cohesion as a mechanism, or mediator, for enhancing community well-being.

The review was undertaken with recognition that cultures around the world, particularly indigenous knowledge systems, have long understood and operationalized understanding of how art participation contributes to social cohesion and well-being in communities (56, 57). This indigenous understanding is articulated in a recent study in Canada, which reported that 53.7% of Indigenous residents surveyed reported that the arts are very important to social connection compared with 23.8% of non-Indigenous respondents. Additionally, 63.9% of Indigenous respondents reported that the arts were very important to community well-being, compared to 30.6% of non-Indigenous respondents (58).

Findings of this review also support a growing body of current evidence that articulates the value of arts participation to both well-being and health (5, 55, 59). Although it was not a focus of this review and therefore not reported as a finding, many of the articles in this review reported health benefits of arts participation that align with this literature.

In linking social cohesion to enhanced well-being, this study aligns with a recent analysis that used data from 374,378 individuals in the European Social Survey and found that social cohesion had a very strong and significant effect on subjective well-being (23). It also aligns with the recent *We-Making* initiative in the US, wherein the authors offered a theory of change - representing findings from a study that encompassed literature review, case studies, interviews, logic-modeling, and a two-day expert convening – that links place-based arts and cultural strategies with social cohesion and equitable community well-being (28).

This review engaged the primary elements of the *We-Making Theory of Change* (See Figure 5) as a framework in its design (28). It did not seek to test this theory of change but did seek to identify studies that articulated similar relationships between its primary components, considering the broader concept of arts participation as an abstraction of “place-based arts and cultural strategies” and generalizing from “equitable community well-being” to community well-being.

**Figure 5:** We-Making Theory of Change, reproduced from Engh et al., 2021, p. 11, courtesy of Metris Arts Consulting.

Notably, the findings of this review align with the concept of equitable community well-being articulated in the *We-Making* model, as included studies commonly reported improvements in mental and physical health (37–54), preservation of culture (40–42, 44, 50, 52), creative responses to trauma and racism (40, 44, 50), and civic capacity for change (38, 40, 44–46, 48, 49, 51, 52, 54). It also aligns with the *We-Making* assertion that “this process feeds back into, amplifies, and grows social cohesion” (p. 11). This review builds on this theory of change with a nuanced understanding of *how* arts programs – with or without an explicit focus on social issues or social change – can build the drivers of social cohesion and the commitments that result in sustained participation and persistent presence in a community.

This review’s articulation of a cycle of community-based arts participation, co-creation and cultural sharing, relationships and belonging, commitment, and more regular arts participation highlights the usefulness of co-creation, in particular, as a key element in arts programs designed to enhance social cohesion and community well-being. Overall, this review supports the *We-Making Theory of Change* and offers this additional insight as well as a more generalized conceptual model for engaging the arts for social cohesion and well-being in communities.

The review was also interested in identifying research wherein, like in the *We-Making Theory of Change*, social cohesion was identified as a mechanism to well-being. Six such studies were found (see results section above), lending support for this theory of change. However, this idea requires further research as some studies have also noted complexities that should be considered. For example, in a study of social capital in participatory arts programs for well-being, the authors explored two key dimensions of social capital – bonding and bridging – and noted that bonding in the absence of sufficient bridging in arts projects can reinforce unequal social relations that are detrimental to health and well-being (60). Notably, some of the variables of social capital explored in that study overlapped with the components of social cohesion used in this review. Additionally, solidarity emerged as a common component of social cohesion in this review and should be investigated further as a potentially important component in building social cohesion.

Importantly, the diverse culturally-rooted participatory arts programs included in this review contribute to understanding of the importance of the arts for preserving culture and for bridging cultures and cultural differences in community groups. Good et al (2021) highlighted how song and dance, drumming, and learning traditional lyrics in a school-based music program enhanced Indigenous youth’s connections with their heritage (40). Additionally, some studies highlighted how other forms of arts participation, such as public-square dancing and community circus arts can create space for exploring attachment to place and land, such as when community arts centers help preserve cultural identity and pride and create gathering space for immigrant communities (50–52).

This review suggests that community gardens and choirs may be particularly useful art forms for building social cohesion and, in turn, well-being in communities. This may be because through membership, choirs can provoke a sense of belonging and inclusion and, through group singing, a sense of unity. Community gardening may provide unique opportunities for building social relationship and networks, trust, and social capital. Both forms include co-creation and regular participation and are activities that tend to be available in many communities, and that many people enjoy and may be easily drawn to as compared to forms such as dance, circus, or magic.

Notably, the findings of this review are reflected in the designs of three past and current nationally scaled arts programs, all intended to address social cohesion and well-being in communities in the US. First, the Federal Art Project component of the Works Progress Administration (WPA) of 1935, implemented in response to the need for rebuilding the economic and social landscape of US communities following the Great Depression (61). The WPA’s unprecedented investment in the arts demonstrated an understanding of the power of the arts to help address social problems at scale.

Secondly, the Public Works program, which engages theatre and the arts to “restore and build community by connecting people through the creation of extraordinary works of art” (62). This program, which has been implemented in communities across the US and beyond, is grounded in the understanding of three things that lead to group transformation - group enterprise, safe boundaries, and high stakes (63). Recognizing that community co-creation of art works meets this definition, the Public Works program implements large-scale community co-creation and staging of theatre works that are reported by participants to yield “significant mental and physical health benefits, along with a deep sense of bonding and social cohesion” (63). Finally, and emanating directly from the inspiration of these two programs, One Nation/One Project is a national initiative in the US designed to engage the arts at a national scale to rebuild the social fabrics and well-being of communities following the COVID-19 pandemic. In this initiative, artists in 18 towns and cities are working with local public health and municipal partners to create large-scale participatory arts projects.

### Strengths, Limitations and Recommendations for Further Research

One strength of this review is its breadth and inclusiveness. Integrative reviews allow for more breadth than systematic reviews or meta-analyses, contributing to development of theory and expansion of scientific thought where topics or questions are under-explored (64, 65). Additionally, by extending from the formative historical work on social cohesion, this review provides a current perspective of its role in relation to community-based arts and community well-being. Despite growing interest in both the arts and social cohesion in relation to well-being, this is the first review to consider these three concepts together. Another strength of this review is its broad definition of arts participation, which allowed inclusion of some practices that previous definitions have excluded, such as circus arts. Additionally, the review supports growing current interest in how the arts can be used for health promotion and prevention (66). Lastly, the prevalence of mixed methods studies in this review provided both quantitative and qualitative perspectives.

This review had several limitations. Firstly, only studies published in English were included, limiting the potential breadth of understanding from communities of non-English speakers, notably Indigenous communities from North America and elsewhere that hold deep knowledge and long-standing practice at the intersections of the arts and health. This review was also limited in its ability to describe the roles of race or ethnicity as a component in relationships between arts participation, social cohesion, and well-being due to inconsistent reporting of race in the included articles. Reporting guidelines for arts programs and interventions in public health would be helpful in such analyses in the future. Additionally, study populations primarily represented urban communities, limiting the applicability of findings to rural communities, and the scale of evidence included in this review does not provide enough evidence to guide replicability of programming, in general.

Future research should include broader representation of ethnic, cultural, and social arts programs and practices in rural communities to inform programming in communities outside of urban areas. Additionally, more quantitative methods and population studies may inform reliability and reproducibility of results, aiding in the confirmation of effects of arts-based practices on social cohesion and well-being. Further study of virtual communities should be considered as well, given that results related to the benefits of virtual communities to social cohesion and well-being were mixed in this review, and given the rise in virtual communities since the pandemic. Future studies should also explicitly test the potential role of social cohesion as a mechanism for building both individual and collective well-being in communities.

## Conclusions

Findings of this integrative review support previous assertions that arts participation may be a useful approach to enhancing social cohesion and well-being in communities. Notably, it identified dimensions that may offer new insight regarding the relationships between arts participation, social cohesion, and well-being. As represented in its conceptual model, the review suggests that community-based arts participation, particularly when co-creation and/or cultural sharing are involved, can help build relationships, a sense of belonging to a community, and solidarity. This can lead to a greater commitment to the program and fellow participants, which in turn can lead to more regular participation in the program. This cycle of arts participation can build social cohesion and (potentially, in turn) enhance both individual and community well-being. Prospective studies are needed to investigate these relationships. The review also suggests that the social cohesion that is cultivated through arts participation may act as a mechanism for enhancing well-being. Further research is needed to explore this relationship. At a time when loneliness and social isolation are of paramount concern in the US and in other parts of the world, this review contributes to understand of how community-based arts participation may build social cohesion and, perhaps in turn, community well-being.

## Data Availability

The data underlying the results presented in the study are available from the University of Florida Center for Arts in Medicine's research landing page (https://arts.ufl.edu/academics/center-for-arts-in-medicine/researchandpublications/). Data will be made available on this landing page upon acceptance by the journal.

https://drive.google.com/file/d/1pu5_Q8OWdVFoc4koqsukyH89bqbJrw_d/view?usp=sharing

## Acknowledgements

The authors would like to acknowledge the contribution of University of Florida’s Center for Arts in Medicine’s Interdisciplinary Research Lab members, Jennifer Kuo, Caroline Wagner, Xander Boggs, Tessa Brinza, and Sohrob Farahbakhsh, as well as Mariana Occhiuzzi, Tyler Thomas, Michael Rohd, and Lear DeBessonet. Funding for One Nation/One Project was provided by Anne Clarke Wolff and Ted Wolff, Barbara and Amos Hostetter, Bloomberg Philanthropies, Create Foundation, Doris Duke Foundation, Frances Clayton & Jessi Hempel, Hull Family Foundation, Jason Cooper, Katie McGrath & J.J. Abrams Family Foundation, Kevin Ryan, The Kresge Foundation, Mortimer & Mimi Levitt Foundation, Lyle Chatelain Family Foundation, Mellon Foundation, The Robert and Mercedes Eichholz Foundation, Sozosei Foundation, and The Tow Foundation.

## Supporting Information Captions

S1 Search Strategy: Sample Search Strategy for PubMed

S2 PRISMA ScR Checklist

