## Supplemental Materials for "Relationships between arts participation, social cohesion, and wellbeing: An integrative review and conceptual model"

### PubMed Search Strategy

| Set Number | PubMed Search Terms |
|---|---|
| #1 | tiab(art OR arts OR artist OR artists OR artistic OR artistry OR quilt* OR "silk screen" OR "silk screened" OR "silk screening" OR acting OR actress* OR playwrit* OR "playwright*" OR jazz OR "art-based" OR "arts-based" OR "creative effort" OR "creative efforts" OR "creative engagement" OR "creative expression" OR "creative expressions" OR "creative process" OR "creative processes" OR "creative medicine" OR "creative practice" OR "creative therapy" OR "creative therapies" OR "creative writing" OR "group writing" OR "entertainment education" OR "expressive activity" OR "expressive activities" OR "expressive writing" OR "graphic novel*" OR "journal writing" OR "diary writing" OR "movement therapy" OR "movement therapies" OR roleplay* OR "role play" OR "role playing" OR "role-play" OR "role-playing" OR "role-plays" OR animat* OR artwork* OR ballet OR caricature* OR cartoon* OR choir* OR choreography OR choreographing OR choreographer* OR clay OR collag* OR comic OR comics OR comicbook* OR danc* OR drama* OR psychodrama* OR edutainment OR film OR films OR filming OR improvisation* OR improvization* OR journaling OR mandala* OR mural* OR museum* OR music* OR novela* OR novella* OR opera OR operas OR operatic OR paint* OR pictorial OR plays OR poet* OR poem* OR haiku* OR portrait* OR potter* OR puppet* OR rap OR raps OR rapping OR sculpt* OR sing OR singing OR sings OR singer* OR skit OR skits OR song* OR story OR stories OR storyline* OR storytell* OR textile* OR theatr* OR theater* OR watercolor* OR "water-color" OR "water-colors" OR "water color" OR "water colors" OR watercolor* OR ceramics OR mosaic* OR lyrics OR graffiti OR "hip hop" OR aesthetic* OR esthetic* OR "spoken word" OR sketch* OR coloring OR movie* OR webcast* OR "motion picture" OR "motion pictures" OR cinema* OR gallery OR galleries OR "narrative therapy" OR "narrative therapies" OR sew OR sews OR sewing OR weave OR weaves OR weaving OR crochet* OR knit OR knits OR knitting OR spinning OR needlework* OR needlepoint* OR macram* OR embroider* OR "rug hooking" OR tapestr* OR dyeing OR "tie-dy*" OR temari OR shibori OR paperfold* OR "paper folding" OR origami OR scrapbook* OR stamping OR collage OR collaging OR collages OR decoupage OR calligraphy OR papercutting OR "paper cutting" OR papercraft* OR paperart* OR papercraft* OR craft OR crafts OR crafting OR quilling OR papermaking OR "paper making" OR printmaking OR marbling OR screenprint* OR "screen printing" OR "paper mache" OR "papier mache" OR illustration* OR whittling OR woodcarving OR carving OR "wood work*" OR coopering OR cooperage OR woodburn* OR pyrography OR inlay OR enameling OR cloisonn* OR engraving* OR embossing* OR etching* OR "wire work*" OR "metal work*" OR metalwork* OR blacksmith* OR smithing OR tinsmith* OR goldsmith* OR silversmith* OR beading OR handbuilding OR "hand building" OR "glass blowing" OR glassblow* OR "lamp work*" OR "stained glass" OR basketmak* OR basketry OR ikebana OR "flower arrang*" OR "floral arrang*" OR "leather work*" OR leatherwork* OR batik* OR lithograph* OR "jewelry mak*" OR jewelrymak* OR stitchery OR handicraft* OR sandplay* OR "sand play*" OR |

### PubMed Search Strategy

| | |
|---|---|
| | sandpaint* OR decorat* OR “culinary practice*” OR “industrial design” OR fiction OR “non-fiction” OR memoir OR memoirs OR screenwriting OR choral OR “stage design*” OR “costume design*” OR “architectural design*” OR gardening OR “creative hobby” OR “creative hobbies” OR bharyamana OR wacking OR vogue OR breakdanc* OR kathak OR odissi OR merengue OR bachata OR salsa OR samba OR rumba OR chacha OR krump OR tango) |
| #2 | ("Art"[Mesh] OR "Sensory Art Therapies"[Mesh] OR "Art Therapy"[Mesh] OR "Dancing"[Mesh] OR "Gardening"[Mesh] OR "Architecture"[Mesh:NoExp] OR "Interior Design and Furnishings"[Mesh] OR "Printing"[Mesh] OR "Printing, Three-Dimensional"[Mesh] OR "Ceramics"[Mesh:NoExp] OR "Poetry as Topic"[Mesh] OR "Webcasts as Topic"[Mesh] OR "Motion Pictures"[Mesh] OR "Hobbies"[Mesh] OR "Music"[Mesh] OR "Music Therapy"[Mesh] OR "Drama"[Mesh] OR "Psychodrama"[Mesh] OR "Television"[Mesh:NoExp] OR "Textiles"[Mesh] OR "Books, Illustrated"[Mesh] OR "Blogging"[Mesh] OR "Color Therapy"[Mesh] OR "Creativity"[Mesh] OR "Diaries as Topic"[Mesh] OR "Graphic Novels as Topic"[Mesh] OR "Portraits as Topic"[Mesh] OR "Narration"[Mesh] OR "Narrative Therapy"[Mesh] OR "Narrative Medicine"[Mesh] OR "Caricatures as Topic"[Mesh] OR "Cartoons as Topic"[Mesh] OR "Imagination"[Mesh:NoExp] OR "Museums"[Mesh] OR "Paintings"[Mesh] OR "Radio"[Mesh] OR "Play Therapy"[Mesh] OR "Singing"[Mesh] OR "Imagery, Psychotherapy"[Mesh] OR "Esthetics"[Mesh] OR "Ceramics"[Mesh] OR "Textiles"[Mesh] OR "Fictional Works as Topic"[Mesh]) |
| #3 | #1 OR #2 |
| #4 | tw(antiretroviral OR "anti retroviral" OR "HAART" OR "assisted reproductive therapy" OR "assisted reproductive therapies" OR "state of the art") |
| #5 | #3 NOT #4 |
| #6 | ti(Participat* OR making OR make OR makes OR maker OR makers OR active OR activity OR activities OR engag* OR listen* OR view* OR watch* OR danc* OR involv* OR partak* OR share OR shares OR sharing OR shared OR association OR contribut* OR concurrence OR attend* OR consuming OR playing OR plays OR sing OR sings OR singing OR singer* OR observer* OR experienc* OR interact* OR program* OR workshop* OR class OR classes OR forum OR forums OR perform* OR group OR groups OR practice OR practicing OR practiced OR hobby OR hobbies OR collab* OR placemak* OR “cultural practice*” OR event OR events OR festival* OR fair OR fairs OR creativ* OR “social citizenship” OR “community action” OR audience* OR exhibit* OR project* OR express* OR attend* OR “art learning” OR “arts learning”) |
| #7 | ("Social Participation"[Mesh] OR "Community Participation"[Mesh] OR "Dancing"[Mesh] OR "Hobbies"[Mesh] OR "Singing"[Mesh]) |
| #8 | #6 OR #7 |
| #9 | tiab(local OR localit* OR neighborhood* OR neighbourhood* OR statewide OR communit* OR communal OR "public health" OR “population health” OR "CBPR" |

### PubMed Search Strategy

| | |
|---|---|
| | OR city OR cities OR municipal* OR county OR counties OR town* OR village* OR suburb* OR region* OR parish* OR diocese* OR district* OR barrio*) |
| #10 | ("Cities"[Mesh] OR "Suburban Population"[Mesh] OR "Urban Population"[Mesh] OR "Poverty Areas"[Mesh] OR "Local Government"[Mesh] OR "Suburban Health Services"[Mesh] OR "Rural Health Services"[Mesh] OR "Population Health"[Mesh]) |
| #11 | #9 OR #10 |
| #12 | tiab(“social cohesi*” OR “community cohesi*” OR “cohesive societ*” OR “cohesive communit*” OR “community ownership” OR “civic engagement” OR belong OR belongs OR belonging OR “social participation” OR “place attachment” OR “social capital” OR “common good” OR “community building” OR “building communit*” OR “community development” OR “community rehabilitation” OR connectedness OR “collective voice*” OR “community resilience” OR “civic capacity” OR “community capacity” OR “social inclusion” OR “community inclusion” OR “community integrat*” OR “social capital” OR “place making” OR “place attachment” OR “community participation” OR “cultural participation” OR “community engagement” OR “social engagement” OR “community connection*” OR “community network*” OR “sense of community” OR “social tolerance” OR altruism OR altruistic OR “civic integrat*” OR “social integrat*” OR “social reintegra*” OR “social activism” OR “community activism” OR “community quality of life” OR “social support” OR “community support” OR “social isolation” OR “socially isolated” OR “social discrimination” OR “social exclusion” OR “social estrangement” OR “cultural exclusion” OR “social breakdown syndrome*” OR “social relations” OR “community solidarity” OR “social solidarity” OR “sociocultural participation” OR “sociocultural cohesi*” OR “sociocultural engagement” OR “sociocultural inclusion” OR “community conversation*” OR “community vitality” OR “social tie” OR “social ties” OR “community tie” OR “community ties” OR “social trust” OR “community trust” OR “density of social relation*” OR “community function*” OR “social identity” OR “community identity” OR “community bond*” OR “social bond*” OR “social connectiv*” OR “community connectivity” OR “social contact” OR “social communication” OR “social exclusion” OR loneliness OR lonely OR ostracis* OR “social alienation” OR “social deprivation” OR “social interaction*” OR “social cooperation” OR “social co-operation” OR “community cooperation” OR “community co-operation” OR “social functioning” OR “social solidarity” OR “community solidarity” OR “cultural participation” OR “social citizenship” OR “community action” OR “community support*” OR “community healing”) |
| #13 | ("Social Isolation"[Mesh] OR "Social Interaction"[Mesh] OR "Social Integration"[Mesh] OR "Social Marginalization"[Mesh]) |
| #14 | #12 OR #13 |
| #15 | tiab(wellbeing OR “well being” OR “well-being”) |
| #16 | #5 AND #8 AND #11 AND #14 AND #15 |
| #17 | #16, ((English[Filter]) AND (2000:2022[pdat])) |

#### Preferred Reporting Items for Systematic reviews and Meta-Analyses extension for Scoping Reviews (PRISMA-ScR) Checklist

| SECTION | ITEM | PRISMA-ScR CHECKLIST ITEM | REPORTED ON PAGE # |
|---|---|---|---|
| **TITLE** | | | |
| Title | 1 | Identify the report as a scoping review. | 1 |
| **ABSTRACT** | | | |
| Structured summary | 2 | Provide a structured summary that includes (as applicable): background, objectives, eligibility criteria, sources of evidence, charting methods, results, and conclusions that relate to the review questions and objectives. | 2 |
| **INTRODUCTION** | | | |
| Rationale | 3 | Describe the rationale for the review in the context of what is already known. Explain why the review questions/objectives lend themselves to a scoping review approach. | 5-6 |
| Objectives | 4 | Provide an explicit statement of the questions and objectives being addressed with reference to their key elements (e.g., population or participants, concepts, and context) or other relevant key elements used to conceptualize the review questions and/or objectives. | 6-7 |
| **METHODS** | | | |
| Protocol and registration | 5 | Indicate whether a review protocol exists; state if and where it can be accessed (e.g., a Web address); and if available, provide registration information, including the registration number. | 8 |
| Eligibility criteria | 6 | Specify characteristics of the sources of evidence used as eligibility criteria (e.g., years considered, language, and publication status), and provide a rationale. | 8-9 |
| Information sources* | 7 | Describe all information sources in the search (e.g., databases with dates of coverage and contact with authors to identify additional sources), as well as the date the most recent search was executed. | 8-9 |
| Search | 8 | Present the full electronic search strategy for at least 1 database, including any limits used, such that it could be repeated. | Supplemental materials-S1 |
| Selection of sources of evidence† | 9 | State the process for selecting sources of evidence (i.e., screening and eligibility) included in the scoping review. | 9-10 |
| Data charting process‡ | 10 | Describe the methods of charting data from the included sources of evidence (e.g., calibrated forms or forms that have been tested by the team before their use, and whether data charting was done independently or in duplicate) and any processes for obtaining and confirming data from investigators. | 10-11 |
| Data items | 11 | List and define all variables for which data were sought and any assumptions and simplifications made. | 10-11 |
| Critical appraisal of individual sources of evidence§ | 12 | If done, provide a rationale for conducting a critical appraisal of included sources of evidence; describe the methods used and how this information was used in any data synthesis (if appropriate). | 10 |
| Synthesis of results | 13 | Describe the methods of handling and summarizing the data that were charted. | 11 |

| SECTION | ITEM | PRISMA-ScR CHECKLIST ITEM | REPORTED ON PAGE # |
|---|---|---|---|
| **RESULTS** | | | |
| Selection of sources of evidence | 14 | Give numbers of sources of evidence screened, assessed for eligibility, and included in the review, with reasons for exclusions at each stage, ideally using a flow diagram. | Figure 1. PRISMA Diagram |
| Characteristics of sources of evidence | 15 | For each source of evidence, present characteristics for which data were charted and provide the citations. | 12-15 |
| Critical appraisal within sources of evidence | 16 | If done, present data on critical appraisal of included sources of evidence (see item 12). | 12-13 |
| Results of individual sources of evidence | 17 | For each included source of evidence, present the relevant data that were charted that relate to the review questions and objectives. | 12-15 |
| Synthesis of results | 18 | Summarize and/or present the charting results as they relate to the review questions and objectives. | 12-15 |
| **DISCUSSION** | | | |
| Summary of evidence | 19 | Summarize the main results (including an overview of concepts, themes, and types of evidence available), link to the review questions and objectives, and consider the relevance to key groups. | 15-23 |
| Limitations | 20 | Discuss the limitations of the scoping review process. | 30-31 |
| Conclusions | 21 | Provide a general interpretation of the results with respect to the review questions and objectives, as well as potential implications and/or next steps. | 31-32 |
| **FUNDING** | | | |
| Funding | 22 | Describe sources of funding for the included sources of evidence, as well as sources of funding for the scoping review. Describe the role of the funders of the scoping review. | 33 |

JBI = Joanna Briggs Institute; PRISMA-ScR = Preferred Reporting Items for Systematic reviews and Meta-Analyses extension for Scoping Reviews.

* Where *sources of evidence* (see second footnote) are compiled from, such as bibliographic databases, social media platforms, and Web sites.

† A more inclusive/heterogeneous term used to account for the different types of evidence or data sources (e.g., quantitative and/or qualitative research, expert opinion, and policy documents) that may be eligible in a scoping review as opposed to only studies. This is not to be confused with *information sources* (see first footnote).

‡ The frameworks by Arksey and O'Malley (6) and Levac and colleagues (7) and the JBI guidance (4, 5) refer to the process of data extraction in a scoping review as data charting.

§ The process of systematically examining research evidence to assess its validity, results, and relevance before using it to inform a decision. This term is used for items 12 and 19 instead of "risk of bias" (which is more applicable to systematic reviews of interventions) to include and acknowledge the various sources of evidence that may be used in a scoping review (e.g., quantitative and/or qualitative research, expert opinion, and policy document).

*From:* Tricco AC, Lillie E, Zarin W, O'Brien KK, Colquhoun H, Levac D, et al. PRISMA Extension for Scoping Reviews (PRISMAScR): Checklist and Explanation. Ann Intern Med. 2018;169:467–473. doi: 10.7326/M18-0850.
